# Reduced brain entropy in migraine with partial restoration during attacks: a resting-state fMRI study

**DOI:** 10.1101/2025.10.29.25339059

**Authors:** Majid Saberi, Dajung J Kim, Xiao-Su Hu, Alexandre F DaSilva

## Abstract

Migraine is a prevalent and disabling neurological disorder, characterized by impaired regulation of migraine burden, sensory processing, and cognitive-emotional states. Brain entropy quantifies the complexity of neural dynamics, where reduced entropy may reflect diminished neural adaptability, but its assessment with fMRI in migraine remains limited. Here, we examined alterations in brain entropy and their associations with clinical burden, migraine phase, and symptomatology. Resting-state fMRI data were acquired from adults with episodic migraine, chronic migraine, and healthy controls. Following standard preprocessing, voxel-wise sample entropy was computed, and group differences were assessed using ANCOVA with age and sex as covariates. Associations with clinical burden and symptom measures were examined within affected regions. In chronic migraine, attack timing-related changes in entropy were further explored, and the Largest Lyapunov Exponent (LLE) was estimated to characterize chaotic dynamics underlying attack-related complexity changes. Migraine patients showed reduced entropy in visual, dorsal attention, and default mode network regions compared to controls, most pronounced in chronic migraine. Lower entropy correlated with greater headache frequency and longer illness duration. In chronic migraine, entropy relatively increased during attacks in multisensory integration regions and was associated with positive and elevated LLEs, indicating partially restored complexity with weakly chaotic dynamics. Patients experiencing phonophobia and nausea also exhibited increased entropy in multisensory integration and default mode network regions. Our findings demonstrate widespread reductions in brain entropy in migraine, reflecting impaired neural adaptability, whereas attacks may transiently restore complexity partially through weakly chaotic dynamics. These results advance understanding of migraine pathophysiology and highlight potential targets for therapeutic intervention.

**Highlights:**

- Migraine is associated with reduced brain entropy across visual, dorsal attention, and default mode network regions, correlating with clinical burden.
- Reduced entropy reflects constrained neural adaptability within affected regions.
- Migraine attacks transiently restore entropy, suggesting partial recovery of neural adaptability.
- Positive and elevated largest Lyapunov exponents indicate a shift toward weakly chaotic dynamics during migraine attacks in multisensory integration regions.
- Symptoms such as phonophobia and nausea are linked to increased entropy in multisensory integration and default mode regions.

## Introduction

Migraine is a prevalent and disabling neurological disorder affecting over one billion individuals worldwide. Between 1990 and 2021, its prevalence rose by 58%, incidence by 42%, and disability-adjusted life years (DALYs) by 58%, underscoring its growing public health burden (Dang et al., 2025). Clinically, migraine is classified as a primary headache disorder characterized by recurrent attacks of moderate to severe headache, often unilateral and pulsating, lasting 4-72 hours, and commonly accompanied by symptoms such as nausea, photophobia, and phonophobia (Headache Classification Committee of the International Headache Society (IHS), 2018). Chronic migraine is defined as ≥15 headache days per month, with ≥8 days meeting migraine criteria, while episodic migraine involves fewer than 15 headache days monthly.

Recent research increasingly conceptualizes migraine not merely as a vascular condition but as a disorder of brain network dysfunction involving widespread disruptions in sensory, affective, and cognitive regulation (Ashina et al., 2021). Functional MRI studies have revealed altered connectivity and neural activations in several large-scale networks, including the default mode network, dorsal attention network, salience network, visual network, and limbic regions. Each of these networks contributes to core migraine symptoms such as sensory hypersensitivity, impaired attention, and emotional and mood disturbances (Zhou et al., 2023; Zhou et al., 2024). Atypical neural functionality in multisensory integration regions such as the superior parietal lobule and superior temporal gyrus further suggests increased sensitivity to negative emotional input and is associated with migraine frequency (Dong et al. 2023; Klamer et al., 2025). Additionally, alterations in functional connectivity across the preictal, ictal, and postictal phases support the characterization of migraine as a cyclic disorder (Esteves et al., 2025).

Despite substantial progress in characterizing large-scale network abnormalities in migraine, most functional MRI research has focused on static measures of brain activity. Recently, a growing body of research has begun to explore the dynamic properties of brain function in migraine, such as time-varying functional connectivity (Lee et al., 2019; Chen et al., 2023) and variability in BOLD signal fluctuations (Lim et al., 2021). However, sample entropy (Richman & Moorman, 2000), a measure of signal complexity reflecting adaptability and regional information processing, has been increasingly applied in fMRI (Wang et al., 2014), but remains rarely investigated in the context of migraine (Wang et al., 2022). This measure has been applied in resting-state fMRI studies of psychiatric disorders such as depression (Liu et al., 2022), anxiety (Fan et al., 2023), schizophrenia (Xue et al., 2019), and ADHD (Sokunbi et al., 2013), as well as neurological disorders including epilepsy (Zhou et al., 2019), multiple sclerosis (Zhou et al., 2016), chronic pain (Del Mauro et al., 2025), and Alzheimer’s disease (Xue et al., 2018), revealing clinically relevant alterations across key networks. Brain entropy analysis could advance understanding of migraine pathophysiology by characterizing the complexity of brain dynamics across regions and phases and may help identify neural markers that bridge mechanistic understanding with potential treatment targets.

While entropy effectively captures the degree of signal complexity, it cannot distinguish whether complexity arises from structured, deterministic (chaotic) dynamics or unstructured, stochastic sources. Chaos theory provides tools that address this distinction (Sprott, 2003). One such measure is the Largest Lyapunov Exponent (LLE), which describes how rapidly two nearly identical system states diverge over time in a reconstructed phase space (Wolf et al., 1985). In simpler terms, it indicates how sensitive a system is to its initial conditions: even tiny differences at the start can lead to increasingly different trajectories. This growing separation reflects dynamical instability, small perturbations are amplified rather than dampened, thereby limiting how far into the future the system’s behavior can be predicted. When such instability occurs in a deterministic system, it is a hallmark of chaotic dynamics.

Taken together, entropy and LLE provide complementary perspectives: entropy captures the overall complexity of a signal, while LLE characterizes the nature of its underlying dynamics (Stam, 2005). For example, simultaneous increases in entropy and LLE are indicative of enhanced deterministic chaos, whereas high entropy with low LLE suggest a greater contribution of stochastic processes. Although LLE has been widely applied in EEG research, particularly in studies of epilepsy and seizure activity (Brari & Belghith, 2022; Yakovleva et al., 2020), its application in fMRI remains limited despite its high potential (Xie et al., 2008; Keilholz et al., 2020). In migraine, where abnormal sensory integration may underlie susceptibility to attacks (Kernick, 2005), LLE could help reveal whether altered brain dynamics reflect deterministic chaotic processes or stochastic variability, providing mechanistic insights beyond what entropy alone can offer.

In this study, we investigated the complexity of resting-state brain activity in individuals with episodic migraine, chronic migraine, and healthy controls using voxel-wise entropy analysis of fMRI data. Our primary aim was to identify brain regions exhibiting altered signal entropy in migraine, expecting these alterations to be amplified with increasing disease severity. We hypothesized that the diagnosis of migraine would be associated with reduced entropy, indicating constrained neural processing and diminished neural adaptability, in multisensory integration and cognitive-emotional cortical systems where flexible functionality is critical.

To capture dynamic changes associated with migraine attacks, we further examined how brain signal complexity varied across the ictal (during headache), post-ictal (soon after headache), and interictal (outside attack) phases by relating entropy of affected regions to the timing of the most recent attack in chronic migraine patients. We also computed the LLE to complement the entropy analysis and further characterize the nonlinear dynamics underlying migraine attacks. This dual-measure approach allows us to determine whether entropy changes reflect a shift toward enhanced deterministic chaos or a rise in stochastic, noise-dominated activity. We hypothesized that migraine attacks would be associated with increased entropy and elevated Lyapunov exponents, consistent with a state of greater dynamical instability and deterministic chaos. This transient shift may disrupt overly rigid neural states and partially restore adaptive processing in affected systems. Finally, we explored the association between entropy in affected regions and specific migraine-related symptoms, providing further insight into how signal complexity may relate to symptom expression in migraine.

Collectively, this work aims to advance understanding of migraine pathophysiology and highlight novel neural targets for therapeutic intervention.

## Methods

### Participants

Participants included adults aged 18-65 years diagnosed with episodic migraine, chronic migraine, or classified as healthy controls. All participants provided written informed consent, and the study received approval from the Institutional Review Board of the University of Michigan Medical School.

The chronic migraine group included individuals meeting ICHD-3β criteria for chronic migraine (Headache Classification Committee of the International Headache Society (IHS), 2018), defined as experiencing headaches on ≥15 days per month for more than 3 months, with ≥8 days per month having migraine features. Additional inclusion criteria included migraine onset before age 50, and a stable medication regimen for at least 3 months, if applicable. The episodic migraine group consisted of individuals experiencing between 1 and 14 migraine days per month for at least 3 months. These participants were recruited as part of an earlier study cohort and were scanned during the interictal period. All participants were required to have no history of opioid use in the past 6 months and no overuse of analgesics (defined as use on ≥15 days per month). Those with any other chronic pain condition, major psychiatric or neurological disorder, substance use, or MRI contraindications were excluded.

The healthy control group included individuals without a history of migraine, chronic pain, or systemic medical or psychiatric conditions. They were not taking any regular medications affecting the central nervous system and had no MRI contraindications.

### Clinical Assessments

To assess migraine burden, participants with a migraine diagnosis reported the average number of headache days per month and the duration of their illness, defined as the number of years since migraine onset, during the baseline clinical intake visit.

At the time of the MRI scan, participants completed a structured Recent Migraine Questionnaire, a study-specific instrument designed to capture the temporal and symptomatic context of their most recent migraine episode. This questionnaire was developed for this study to characterize migraine phase and symptoms in proximity to scanning and included items on the time since last migraine attack (e.g., current, within the past 12/24/48 hours, or more than 48 hours ago) and presence of symptoms such as photophobia, phonophobia, and nausea/vomiting. This instrument was administered only to the chronic migraine cohort; episodic migraine participants were recruited in an earlier cohort in which recent migraine timing and ictal/interictal state were not systematically collected.

### MRI Data Acquisition

All imaging data were collected on a GE Discovery MR750 3T scanner using a Nova 32-channel head coil. The protocol included resting-state fMRI (rs-fMRI), high-resolution T1-weighted structural scans, and dual-echo fieldmaps for B0 distortion correction.

Resting-state fMRI data were acquired using a multiband EPI sequence (TR = 800 ms, TE = 30 ms, flip angle = 52°, voxel size = 2.4 mm³, 90 × 90 matrix, multiband factor = 6, duration ≈ 6 min). Participants kept their eyes open, fixated on a cross, and minimized movement. T1-weighted images were acquired using a 3D BRAVO sequence (TR = 12.2 ms, TE = 5.2 ms, TI = 500 ms, flip angle = 15°, voxel size = 1 mm³) for anatomical normalization and coregistration. Fieldmaps were collected using dual-echo gradient-echo scans with opposing phase encoding (AP/PA), TR = 7.4 ms, TE = 80 ms, flip angle = 90°, voxel size = 1.687 × 1.687 × 2.4 mm³. These were used to compute voxel displacement maps for EPI distortion correction.

### Preprocessing of fMRI Data

Resting-state fMRI data were preprocessed using FSL (Jenkinson et al., 2012), fMRIPrep (Esteban et al., 2019), and AFNI (Cox, 1996). First, all images were converted to BIDS format (Gorgolewski et al., 2016). To correct for susceptibility-induced distortions, B0 distortion correction was performed using the FSL TOPUP algorithm, based on dual-echo fieldmaps acquired with opposing phase-encoding directions.

Following distortion correction, data were preprocessed using fMRIPrep, which included head motion correction, co-registration to the T1-weighted structural image, spatial normalization to MNI152 standard space, and brain segmentation. As part of this pipeline, nuisance components, including principal components of white matter and cerebrospinal fluid (aCompCor (Behzadi et al., 2007)) and six motion parameters, were automatically extracted for denoising.

Next, additional denoising was performed using FSL commands to remove the six motion parameters and aCompCor components. The denoised data were then temporally filtered using AFNI commands, applying a bandpass filter from 0.01 to 0.1 Hz to retain biologically relevant low-frequency resting-state fluctuations. Finally, spatial smoothing was applied using FSL, with a Gaussian kernel of 5 mm full width at half maximum (FWHM).

All preprocessing steps were visually checked to ensure data quality and alignment.

### Entropy Computation

To quantify the complexity of resting-state brain dynamics, we computed voxel-wise sample entropy maps from the final smoothed 4D fMRI data (Richman & Moorman, 2000; Wang et al, 2014). For each participant, this process yielded a single 3D entropy map representing local signal irregularity across the brain. Sample entropy was computed using a custom implementation in R with embedded C++ code for efficient processing.

For each voxel, the time series was extracted and entropy was calculated using an embedding dimension (m) of 2 and a similarity threshold (r) equal to 20% of the standard deviation of the voxel’s time series. Entropy computation was restricted to voxels within a binarized MNI152 brain mask to reduce computational load and exclude non-brain regions. These individual entropy maps were subsequently used for voxel-wise group-level statistical analyses to identify regional differences in signal complexity across study groups. In this framework, lower entropy indicates more constrained neural dynamics, while higher entropy reflects more adaptable neural dynamics.

To evaluate the robustness of entropy estimates to parameter selection, additional entropy maps were computed using alternative parameter configurations. Specifically, entropy was recalculated using an increased embedding dimension (m = 3, r = 0.2 × SD) and an increased similarity tolerance (m = 2, r = 0.25 × SD). These alternative entropy maps were used in subsequent sensitivity analyses to assess the stability of entropy estimates across parameter settings and to confirm that the observed entropy patterns were not dependent on specific parameter choices.

### Lyapunov Analysis

To further investigate the nonlinear dynamical characteristics underlying changes in signal complexity in affected regions, we examined the LLE of their mean time series. The LLE quantifies the average rate at which nearby trajectories in a reconstructed phase space diverge over time, serving as a marker of deterministic chaos by capturing sensitivity to initial conditions, as opposed to stochastic variability (Wolf et al., 1985; Rosenstein et al., 1993).

The estimation of the LLE follows a previously published method (Xu et al., 2011). Time-delay embedding was used to reconstruct phase space dynamics from the fMRI signal. The optimal time delay and embedding dimension were determined using established methods: average mutual information for delay selection and the false nearest neighbors algorithm to identify the appropriate embedding dimension. To minimize the influence of autocorrelation, a Theiler window was applied based on the signal’s dominant frequency component.

LLE values were then computed using the algorithm which estimates divergence over time between neighboring trajectories (Rosenstein et al., 1993). This approach is robust to noise and particularly well-suited for short physiological time series like those in fMRI. In this framework, positive LLE values indicate divergence and dynamical instability (consistent with chaos), values near zero indicate neutral or marginally stable dynamics, and negative values indicate convergence toward stable attractors. Given the technical limitations of short and noisy fMRI time series, the magnitude of LLE values is typically small; therefore, in this study we interpret positive but low values as indicative of weakly chaotic behavior rather than strong deterministic chaos (Bunimovich & Su, 2025; Heiligenthal et al., 2011).

### Statistical Analysis

Voxel-wise group comparisons of entropy were conducted using analysis of covariance (ANCOVA), with group (chronic migraine, episodic migraine, healthy controls) as the main factor and age and sex as covariates. To correct for multiple comparisons, a cluster-wise threshold was applied to voxels with p < 0.05. Spatial smoothness of model residuals was estimated using AFNI’s 3dFWHMx with the autocorrelation function (ACF) model. Cluster-level family-wise error (FWE) correction was subsequently performed using AFNI’s 3dClustSim based on the estimated ACF parameters, applying two-sided inference with nearest-neighbor connectivity (NN = 3). Residuals from the voxel-wise models were used with AFNI’s tools to estimate a minimum cluster size for statistical significance. Clusters were defined using a 26-voxel connectivity criterion, where voxels were considered contiguous if they shared a face, edge, or corner, to control the family-wise error rate (FWE) at p < 0.05. The resulting cluster-extent threshold corresponded to a minimum of 187 contiguous voxels. Four clusters exceeded the threshold and were identified using FSL tools for subsequent analyses.

To assess robustness to parametric assumptions and the non-Gaussian properties of entropy measures, voxel-wise nonparametric ANCOVA was additionally performed using the Freedman-Lane permutation approach with 5,000 permutations, controlling for age and sex. The spatial distribution of significant clusters derived from the permutation-based analysis was compared with those obtained from the parametric ANCOVA.

To evaluate robustness to preprocessing and denoising choices, the entire preprocessing and group-level entropy analysis was repeated using ICA-AROMA as an alternative denoising strategy. The resulting statistical maps were compared with those obtained using the primary pipeline. In addition, voxelwise temporal signal-to-noise ratio (tSNR) maps were computed for each subject following both denoising approaches, and group-level comparisons were performed to assess differences in signal robustness between pipelines.

To assess potential confounding effects of head motion on the brain entropy, framewise displacement (FD) and DVARS were computed for each participant and compared across groups using one-way ANOVA.

To further assess robustness to entropy computation parameters, the voxelwise group-level analyses were repeated using entropy maps derived from alternative parameter configurations, and the resulting spatial patterns were compared with those obtained using the primary parameter settings.

To assess the potential influence of depressive symptom severity, an additional sensitivity analysis was performed in a subset of participants with available Beck Depression Inventory (BDI) data. Voxel-wise ANCOVA was repeated including BDI score as an additional covariate alongside age and sex. The resulting statistical maps were compared with those obtained from the primary model.

Voxelwise ANCOVA constituted the primary inferential analysis for identifying regions exhibiting group-level entropy differences. Mean entropy values were extracted from significant clusters and used in between-group comparisons and correlations with clinical measures such as headache days per month and illness duration.

To examine transient changes in signal complexity related to migraine attacks, participants with chronic migraine were categorized based on the time since their most recent attack: ictal, <12 hours, 12-24 hours, 24-48 hours, and >48 hours (interictal). Spearman’s rank correlations were performed between entropy values and time since attack, and between LLEs and time since attack in the identified clusters.

To determine whether altered entropy during migraine attacks reflects deterministic chaos or stochastic dominance, we evaluated the relationship between LLE and entropy values in identified clusters using Spearman correlation. A strong positive correlation would support a state of deterministic chaos, whereas a weak or negative correlation would suggest entropy is primarily driven by stochastic processes.

For symptom-based analyses, independent-samples t-tests were conducted to compare entropy values between participants with chronic migraine who did versus did not report phonophobia, nausea, or vomiting during their most recent attack.

For association analyses between entropy and clinical measures, false discovery rate (FDR) correction was applied across tests. Analyses examining migraine phase dynamics and symptom-based subgroup differences were conducted as secondary exploratory investigations within regions identified by the voxelwise analysis; given the limited sample size in chronic migraine, these analyses were not subjected to additional multiple-comparison correction and should be interpreted as preliminary.

Statistical analyses were performed using R (R Core Team, 2025) and relevant packages, including Rcpp (Eddelbuettel & François, 2011) for computational efficiency in entropy computation, and oro.nifti (Whitcher et al., 2011) for neuroimaging data handling. Visualizations were generated with ggplot2 (Wickham, 2016) for statistical plots and MRIcronGL (Rorden, 2025) for brain maps.

## Results

### Demographic and Clinical Characteristics

The final sample included 66 participants: 25 with episodic migraine (EM), 15 with chronic migraine (CM), and 24 healthy controls (HC). Mean age was 41.7 ± 14.4 years in CM, 30.2 ± 11.5 in EM, and 36.1 ± 16.1 in HC. The proportion of females was 93% in CM, 88% in EM, and 87% in HC. CM participants reported 18.3 ± 4.6 headache days per month and an illness duration of 27.4 ± 15.1 years; EM participants reported 5.5 ± 3 headache days and 12.9 ± 8.6 years of illness. Symptom data were only collected systematically in the CM group during the scanning session; among CM participants, 64% reported photophobia, 21% phonophobia, and 43% nausea or vomiting. In total, 43% of CM participants were scanned during an ictal period and all EM participants were scanned during the interictal period. Beck Depression Inventory (BDI) scores were available for a subset of participants (HC: n = 21, EM: n = 25, CM: n = 11), with mean values of 1.58 ± 3.05 in HC, 2.96 ± 2.9 in EM, and 5.57 ± 6.29 in CM. Demographic and clinical characteristics are summarized in Table 1.

**Table 1.**
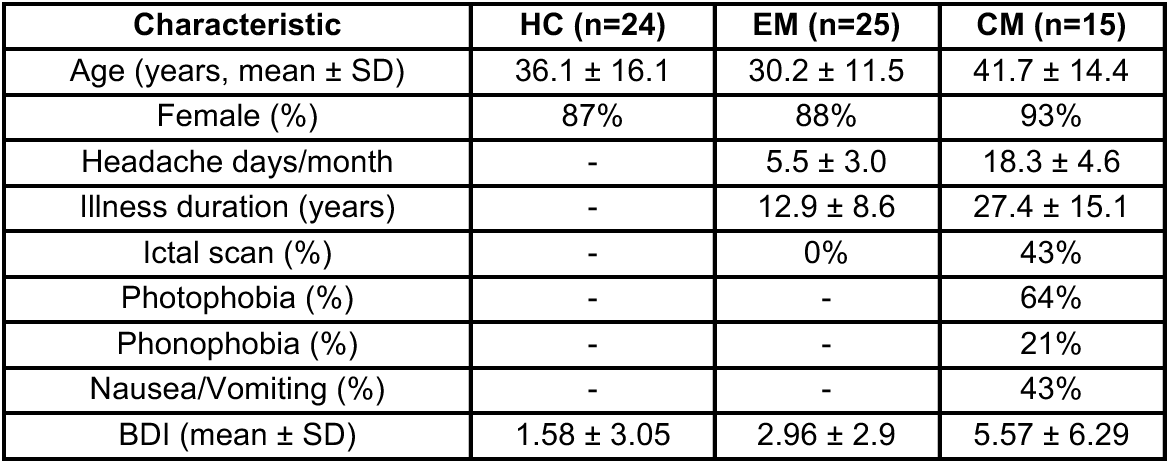
Demographic and clinical characteristics of study participant. (Abbreviations: CM, chronic migraine; EM, episodic migraine; HC, healthy controls; BDI, Beck Depression Inventory.)

### Entropy Decreases in Visual, Dorsal Attention, and Default Mode Networks in Migraine

Four distinct clusters exhibited altered entropy (all surviving cluster-level FWE correction, Figure 1 and Table 2): the occipital cortex (OC), right supramarginal gyrus and superior parietal lobule (rSMG+rSPL), precuneus and posterior cingulate cortex (PCu+PCC), and medial prefrontal cortex (mPFC). In each of these regions, entropy was significantly reduced in the migraine groups compared to healthy controls, with the most pronounced reductions observed in the chronic migraine group (peak voxel statistics are reported in Table 2).

**Figure 1.**
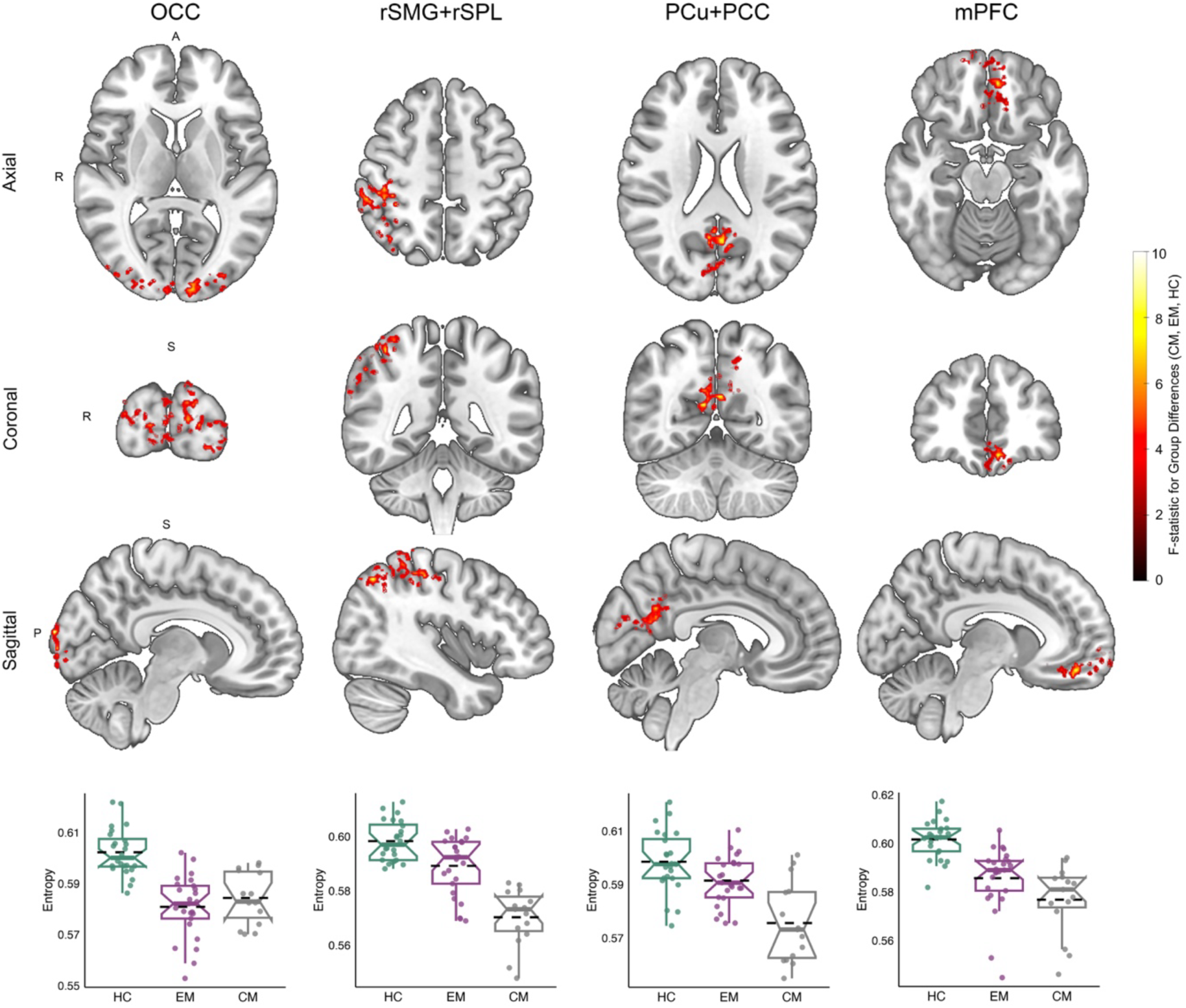
Group-level differences in regional brain entropy. Colored voxels in brain maps indicate group-level significant differences in signal entropy among chronic migraine, episodic migraine, and healthy control subjects. The color bar represents F-statistics from the analysis of covariance (ANCOVA). Voxels have passed correction for false positives and are categorized into four distinct regions, displayed in three viewing orientations. For each region, the lower row displays the distribution of entropy values across groups. Horizontal dashed lines represent group-level entropy averages, while box plots indicate medians, interquartile ranges and confidence intervals. Box plots are presented to illustrate the directionality of group differences identified in the voxelwise analysis. (Abbreviations: OC, occipital cortex; rSMG+rSPL, right supramarginal gyrus and right superior parietal lobule; PCu+PCC, precuneus and posterior cingulate cortex; mPFC, medial prefrontal cortex; CM, chronic migraine; EM, episodic migraine; HC, healthy controls.)

**Table 2.**
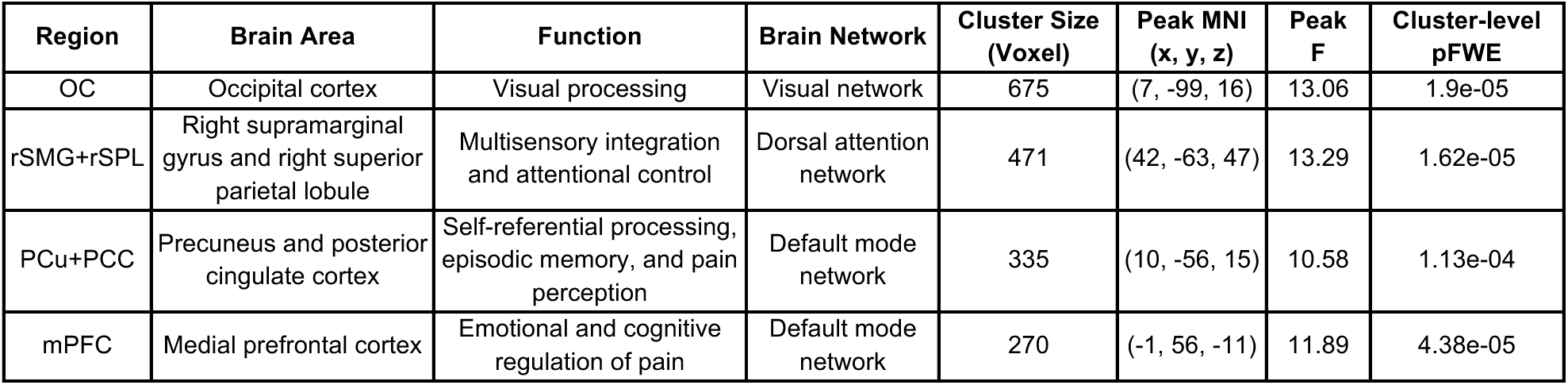
Brain regions showing significantly reduced entropy in migraine patients compared to healthy controls.

Specifically, in the PCu+PCC regions, individuals with chronic migraine showed significantly lower entropy than healthy controls, while those with episodic migraine showed a similar trend that did not reach significance. Chronic migraine patients also had significantly lower entropy than episodic migraine patients in both the PCu+PCC and rSMG+rSPL regions, with a similar trend observed in the mPFC. These graded entropy reductions are consistent with greater disease burden and may reflect increasing constraint in neural dynamics in key visual, dorsal attention, and default mode regions.

To assess robustness and reproducibility, we performed a series of supplementary analyses. These findings were confirmed using a nonparametric permutation-based ANCOVA, which yielded highly similar spatial patterns of group differences (Supplementary Figure S1), supporting the robustness of the results to distributional assumptions.

The spatial pattern of entropy reductions remained largely consistent when alternative denoising strategies were applied (Supplementary Figure S2), and voxelwise signal-to-noise ratio comparisons indicated that the primary preprocessing pipeline provided robust signal quality across the brain (Supplementary Figure S3). In addition, no significant group differences in head motion metrics (framewise displacement and DVARS) were observed (Supplementary Figure S4), suggesting that the findings are unlikely to be driven by motion-related artifacts.

Importantly, sensitivity analyses further demonstrated that the main findings were stable across variations in entropy parameter selection. When using an embedding dimension of m = 3 (Supplementary Figure S5), significant clusters remained in the occipital cortex (OC), rSMG+rSPL, and mPFC, while PCu+PCC showed subthreshold but spatially consistent effects. When increasing similarity tolerance to r = 0.25 × SD (Supplementary Figure S6), significant clusters were preserved in OC, rSMG+rSPL, and PCu+PCC, while mPFC showed subthreshold but consistent effects. These results indicate that the primary spatial pattern of entropy alterations is robust to parameter selection.

Finally, inclusion of BDI as an additional covariate did not substantially alter the main findings, with the major clusters remaining significant (Supplementary Figure S7), indicating consistency of the results after adjustment for depressive symptom severity.

### Reduced Brain Entropy Correlates with Migraine Burden and Illness Duration

Reduced entropy was significantly associated with migraine-related clinical measures across the most regions identified in Figure 1. As shown in Figure 2A, lower entropy values were significantly associated with greater number of headache days per month, particularly in the rSMG+rSPL (r = –0.46, corrected p = 0.01) and PCu+PCC (r = –0.36, corrected p = 0.063; a trend-level after correction). Additionally, longer illness duration (years with migraine) was significantly associated with lower entropy in most regions, including the rSMG+rSPL (r = –0.45, corrected p = 0.008), PCu+PCC (r = –0.49, corrected p = 0.005), and mPFC (r = –0.5, corrected p = 0.005) (Figure 2B). These findings suggest that chronic exposure to migraine is associated with more constrained regional neural dynamics.

**Figure 2.**
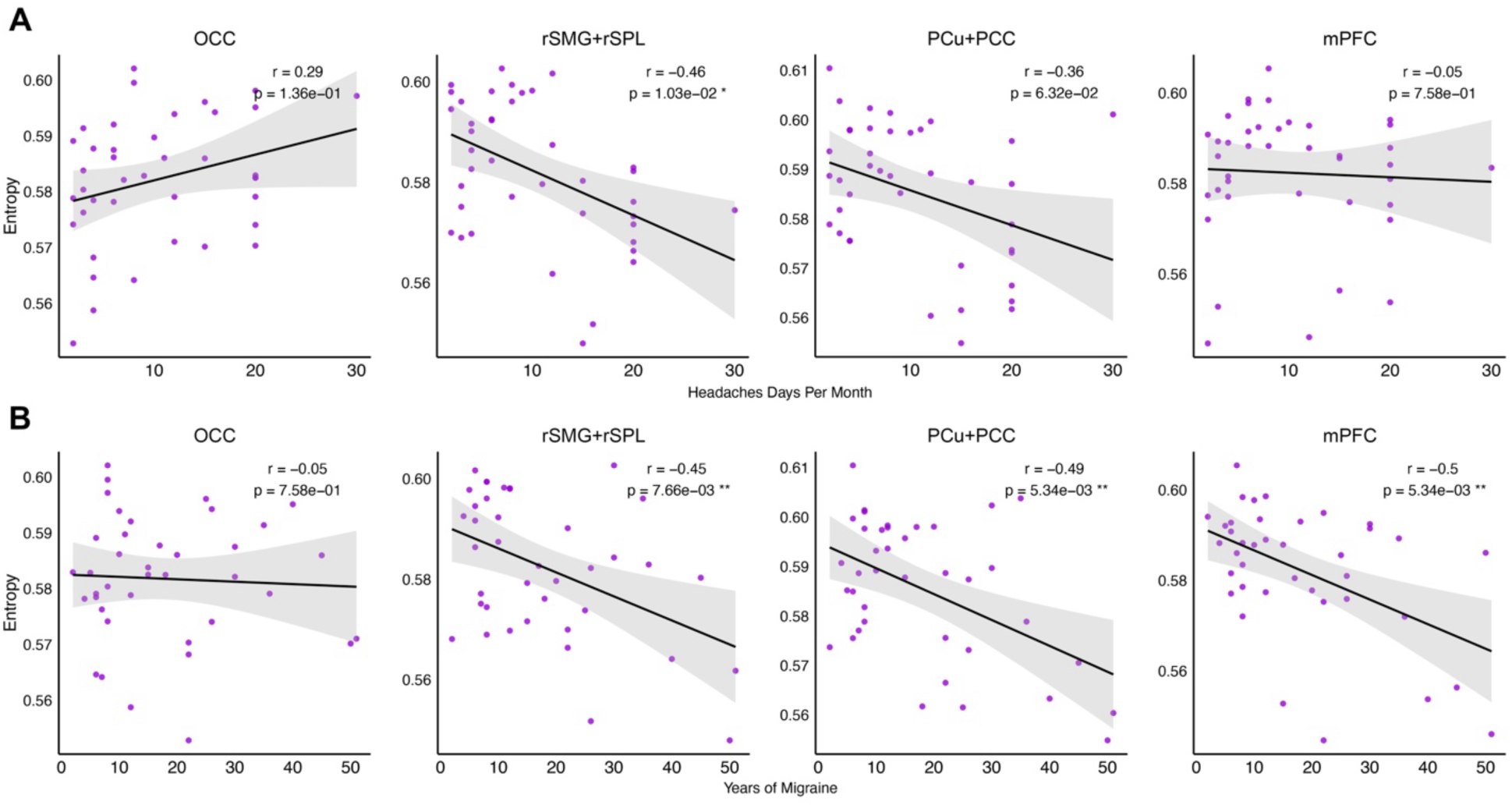
Correlation analysis between brain entropy and general migraine measures across four brain regions with reduced entropy. Each column corresponds to a specific brain region, while the rows represent correlations between brain entropy and headache days per month **(A)**, and brain entropy and years of migraine **(B)**. Each panel displays a scatter plot with a fitted regression line based on data from individuals with migraine. Pearson’s correlation coefficient (r) and the corresponding corrected p-value are reported, with significant correlations marked by an asterisk. (Abbreviations: OC, occipital cortex; rSMG+rSPL, right supramarginal gyrus and right superior parietal lobule; PCu+PCC, precuneus and posterior cingulate cortex; mPFC, medial prefrontal cortex.)

### Migraine Attacks Transiently Elevate Brain Signal Entropy in Key Regions

In chronic migraine, brain entropy varied by time since the most recent attack. Entropy was relatively higher during and shortly after migraine attacks, particularly in rSMG+rSPL (r = –0.55, p = 0.043) and PCu+PCC (r = –0.57, p = 0.034) (Figure 3). Although not significant, similar trends were noted in the OC (r = –0.37, p = 0.19) and mPFC (r = –0.20, p = 0.48). This pattern suggests that although the entropy levels are generally lower in individuals with migraine (inside or outside attacks), migraine attacks temporarily increase brain signal complexity, particularly in parietal and default mode regions involved in multisensory integration, self-referential thought and pain perception, which may reflect a temporary release from the constrained neural state of migraine.

**Figure 3.**
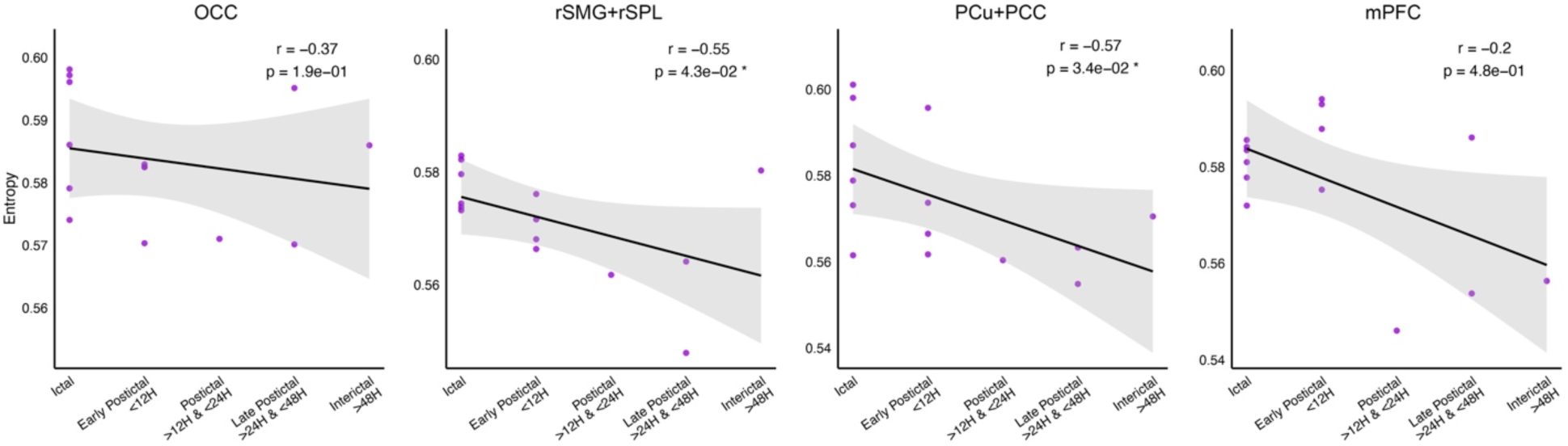
Correlation analysis between brain entropy and the time since the last migraine attack in individuals with chronic migraine. Each column corresponds to a specific brain region that showed altered entropy compared to healthy controls. The x-axis shows categorized time intervals since the last migraine attack (migraine period), while the y-axis represents regional brain entropy. Each panel displays individual data points, a fitted regression line, and the 95% confidence interval. Spearman’s correlation coefficient (r) and the corresponding p-value are reported, with significant correlations (p < 0.05) marked by an asterisk. (Abbreviations: OC, occipital cortex; rSMG+rSPL, right supramarginal gyrus and right superior parietal lobule; PCu+PCC, precuneus and posterior cingulate cortex; mPFC, medial prefrontal cortex.)

### Weakly Chaotic Dynamics Underlie Transient Increases in Brain Entropy During Migraine Attacks

In chronic migraine, Largest Lyapunov Exponent (LLE) values in the rSMG+rSPL were negatively correlated with time since last attack (r = –0.66, p = 0.01), indicating greater dynamic instability near the time of attack (Figure 4A). A marginal positive correlation was observed between LLE and entropy in this region (r = 0.50, p = 0.057), suggesting that entropy increases may reflect underlying chaos rather than pure noise (Figure 4B). Critically, LLE values in this cluster were significantly greater than zero (one-sample t-test vs. 0: t = 5.67, p = 5.75e-05), confirming the presence of diverging trajectories consistent with weakly chaotic-like behavior or dynamical instability rather than strong deterministic chaos. However, the small magnitude of these exponents (<0.1), indicates this is a state of weak chaos or subtle nonlinear regime.

**Figure 4.**
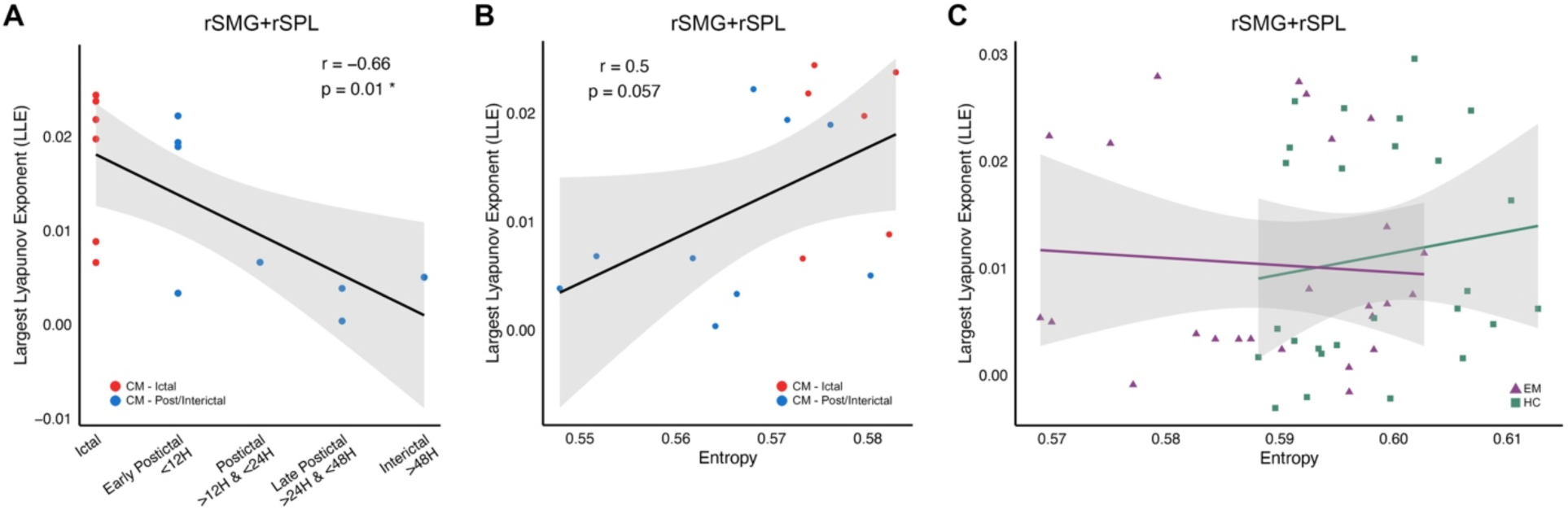
Association between Largest Lyapunov Exponent (LLE), migraine period, and brain entropy in rSMG+rSPL region. **(A)** Correlation between LLE and time since the last migraine attack in chronic migraine. The x-axis shows categorized time intervals since the most recent attack, while the y-axis represents LLE values. **(B)** Correlation between brain entropy and LLE in chronic migraine. Each dot represents an individual, with red dots indicating patients who were in the ictal period of migraine at the time of MRI assessment. **(C)** Correlation between brain entropy and LLE across individuals with episodic migraine and health controls, discriminated by colors. Each panel includes a fitted regression line with 95% confidence intervals. Spearman’s correlation coefficient (r) and the corresponding p-value are reported, with significant correlations (p < 0.05) marked by an asterisk. (Abbreviations: rSMG+rSPL, right supramarginal gyrus and right superior parietal lobule; CM, chronic migraine; EM, episodic migraine; HC, healthy controls.)

No such correlation between entropy and LLE was found in controls or episodic migraine participants scanned during the interictal period (Figure 4C), supporting the specificity of this weakly chaotic state to the attack phase in chronic migraine. These findings suggest that transient disruption of rigid neural states during migraine attacks is accompanied by subtle nonlinear dynamical instability.

### Migraine-Related Symptoms Are Associated with Elevated Entropy in Multisensory Integration and Default Mode Regions

Although chronic migraine patients generally showed reduced entropy, those who experienced phonophobia in recent attack showed relatively higher entropy in rSMG+rSPL (t = 2.51, p = 0.034) (Figure 5A), while those reporting nausea or vomiting showed relatively higher entropy in the PCu+PCC (t = 2.67, p = 0.026) (Figure 5B). The rSMG+rSPL is a key hub for multisensory integration and attentional reorienting, and increased entropy in this region may reflect enhanced neural reactivity in response to sound sensitivity during migraine. The PCu+PCC is part of the default mode network, involved in self-referential thought and interoception. Altered internal state processing or disrupted homeostatic regulation associated with visceral symptoms such as nausea may contribute to elevated entropy in this region.

**Figure 5.**
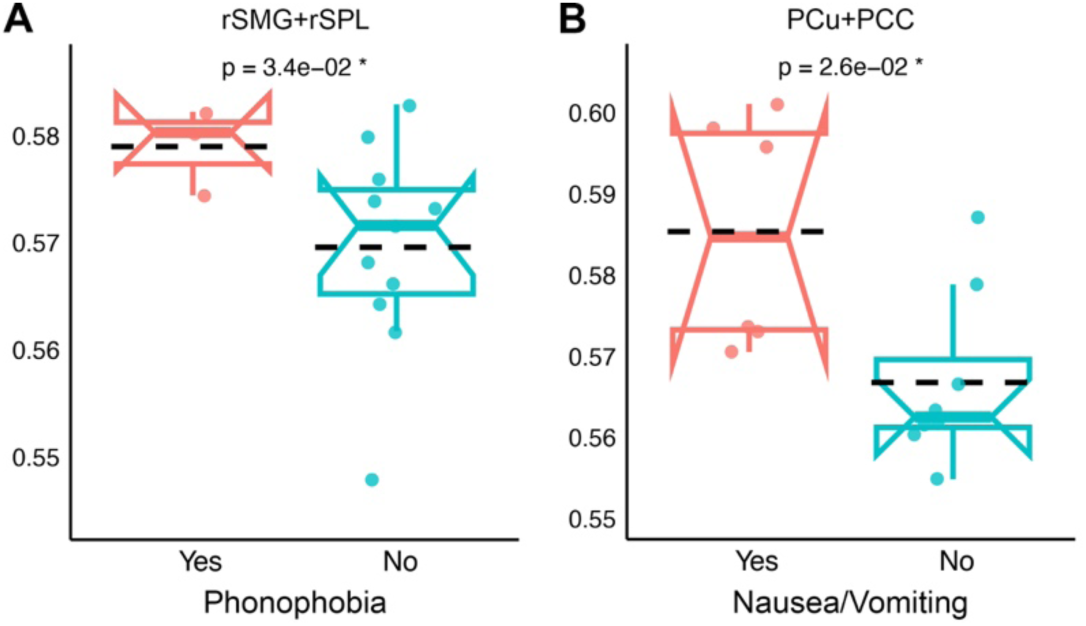
Group differences in brain entropy based on migraine-associated symptoms across individuals with chronic migraine. (A) Comparison of brain entropy in the rSMG+rSPL region between individuals with and without phonophobia. (B) Comparison of brain entropy in the PCu+PCC region between individuals with and without nausea/vomiting. Entropy values are represented by notched box plots showing individual data points, group means (dashed lines), medians, interquartile ranges and confidence intervals. P-values from group comparisons are shown above each panel, with significant differences (p < 0.05) marked by an asterisk. (Abbreviations: rSMG+rSPL, right supramarginal gyrus and right superior parietal lobule; PCu+PCC, precuneus and posterior cingulate cortex.)

## Discussion

### Summary of Findings

In this study, we investigated local brain signal complexity in individuals with episodic migraine, chronic migraine, and healthy controls using voxel-wise entropy and Lyapunov exponent analyses of resting-state fMRI data. Our findings reveal that migraine is associated with widespread reductions in brain entropy, particularly in regions involved in multisensory integration, visual processing, attentional control, self-referential functions, and affective regulation. These reductions were most pronounced in the chronic migraine group and were related to clinical burden, suggesting a progressive constraint in neural adaptability with increasing disease severity. Importantly, we found that entropy values were transiently elevated closer to the time of a migraine attack, the ictal phase, especially in the multisensory integration region, indicating a short-term and partial restoration of the reduced entropy triggered by the attack. The transient ictal increase was further characterized by elevated Largest Lyapunov Exponent (LLE) values, providing evidence that the observed complexity changes reflect underlying weakly chaotic dynamics rather than purely stochastic noise. This suggests that by inducing weakly chaotic dynamics, migraine attacks destabilize rigid neural states and potentially reset adaptive neural processing. Additionally, patients who experienced phonophobia showed increased entropy in multisensory integration areas, while those reporting nausea or vomiting exhibited greater entropy in default mode regions, supporting a link between symptom expression and disruption of migraineurs rigid neural processing. The phase dynamics and symptom-related analyses were conducted as secondary exploratory investigations without additional multiple-comparison correction and should therefore be considered preliminary. These findings collectively highlight the association between regional brain complexity and migraine burden, symptom expression, and migraine phase.

### Cross-Modal Support from EEG Studies

Our findings align with an EEG study that reported reduced entropy during the interictal phase in migraine compared to controls (Cao et al., 2017). Their reductions were limited to the prefrontal cortex, with no changes in occipital regions. In partial agreement, our fMRI results showed reduced entropy in the medial prefrontal cortex but also in occipital regions, contrasting with their findings. We further identified reductions in areas linked to multisensory integration, attention, self-referential, and pain-related processing. While EEG captures rapid scalp-level fluctuations, our voxel-wise fMRI analysis offers higher spatial resolution. These cross-modal findings underscore the value of signal complexity measures in tracking the spatial and temporal dynamics of migraine.

### Comparisons with fMRI-Based Entropy Studies in Migraine

Although we did not differentiate migraine with and without aura, our findings both align with and diverge from a recent fMRI study on patients without aura (Wang et al., 2022). That study used multiscale entropy and reported increased complexity across several networks, including the default mode, frontoparietal, and sensorimotor, depending on temporal scale, with reduced entropy in the posterior DMN at the slowest scale. In contrast, we observed broadly reduced entropy, especially in association with migraine chronicity. Notably, their analysis lacked appropriate correction for multiple comparisons and did not account for covariate effects of age and sex, despite known effects of these factors on entropy (Wang, 2021). These methodological and sample differences may explain the discrepancies. Nonetheless, both studies support altered brain signal entropy in networks linked to self-referential processing, multisensory integration, and attentional control in migraine.

### Parallels with Epilepsy and Shared Entropy Mechanisms

Because both migraine and epilepsy are characterized by episodic alterations in cortical excitability and synchronization, comparisons between them may reveal shared mechanisms of disrupted neural dynamics. Our observation of reduced brain entropy in migraine aligns with epilepsy studies showing decreased neural complexity during interictal periods. Intracranial EEG has consistently demonstrated lower entropy in epileptogenic versus non-epileptogenic regions (Protzner & Valiante, 2010), and gamma-band analyses suggest this reflects excessive neural regularity due to synchronized inhibitory activity (Sato et al., 2019). A resting-state fMRI study in right temporal lobe epilepsy also reported interictal entropy reductions, particularly in attention and sensory integration areas (Zhou et al., 2019). These cross-condition findings suggest that reduced brain entropy may reflect region-specific disruptions in neural adaptability that are common to episodic neurological disorders.

Seizure studies further show that ictal periods involve transient increases in entropy and complexity, with elevated values observed across EEG measures such as distribution entropy, sample entropy, and Lempel–Ziv complexity (Li et al., 2015; Jouny & Bergey, 2012). Similarly, we found increased entropy during migraine attacks, especially in sensory integration regions, suggesting that such paroxysmal events may temporarily disrupt rigid neural states. This supports the idea that both seizures and migraine attacks can transiently restore brain complexity as a compensatory mechanism (Zhou et al., 2016; Zhou et al., 2019).

### Chaotic Dynamics During Migraine Attacks

In addition to entropy, our analysis of Lyapunov exponents offers deeper insight into the nature of increased signal complexity during migraine attacks. LLE values rose in multisensory integration regions near the time of attack, indicating nonlinear dynamical instability. This contrasts with seizure studies, where LLE typically decreases during ictal phases (Khoa et al., 2012).

Given the inherently low signal-to-noise ratio of fMRI BOLD signals, their relatively short temporal length, and the influence of physiological processes such as neurovascular coupling, and because the LLE values we obtained were small and close to zero, we interpret these findings cautiously as evidence for weakly chaotic-like or dynamically unstable regimes rather than definitive proof of strong deterministic chaos.

Importantly, the systematic temporal modulation of LLE in relation to migraine attacks and its association with entropy increases suggest a structured dynamical process rather than purely noise-driven randomness. These results imply that migraine attacks may release the brain from rigid states through weakly chaotic destabilization, enabling escape from pathological attractors.

### Symptom-Related Brain Entropy Patterns

Our results demonstrate that individuals with chronic migraine who experienced phonophobia during their most recent attack showed higher entropy in the right supramarginal gyrus and superior parietal lobule, key multisensory integration regions. This aligns with evidence that overlapping sensory hypersensitivities contribute to migraine-related disability and highlight the role of multisensory processing in migraine (Suzuki et al., 2021; Schwedt, 2013). Structural and functional changes in these regions have also been linked to hypersensitivity and migraine attacks (Chong et al., 2016; Zhang et al., 2022). Our observation of increased entropy associated with phonophobia in these multisensory hubs is consistent with the notion that the migraine brain exhibits abnormal multisensory processing.

We also observed increased entropy in the precuneus and posterior cingulate cortex among individuals reporting nausea or vomiting. These regions are involved in interoception and self-referential processing, and their function has been linked to nausea (Farmer et al., 2015; Ruffle et al., 2019). Our findings suggest that increased signal entropy in default mode regions may reflect disrupted homeostatic regulation during migraine-associated nausea.

### Limitations and Future Directions

This study has some limitations that should be considered. First, its cross-sectional design limits inference about dynamic changes across the migraine cycle, including the preictal phase. Second, the Recent Migraine Questionnaire was administered only in the chronic migraine cohort, which restricts direct comparison of attack-related effects between groups. Third, migraine with and without aura were analyzed together, meaning potential subtype-specific differences could not be addressed in the present dataset. Fourth, the sample size was modest and group sizes were imbalanced across migraine subtypes, which may limit statistical power and generalizability and introduce potential bias, despite the consistency of findings across multiple robustness and sensitivity analyses. Additionally, comprehensive psychiatric assessments, including measures of anxiety and depression, were not available for all participants and therefore could not be consistently incorporated as covariates across the full cohort. As a result, the potential influence of comorbid affective factors cannot be fully excluded.

Future research should employ longitudinal or time-locked designs to track preictal, ictal, and postictal transitions in larger and more balanced migraine samples, enabling a clearer understanding of dynamic changes across the migraine cycle. Incorporating comprehensive psychiatric assessments and larger cohorts will be important to further disentangle migraine-specific neural dynamics from potential comorbid influences. Neuromodulation studies targeting regions with altered complexity may help determine whether restoring neural regulation can reduce migraine burden. Finally, stratifying patients by aura subtype, hormonal status, or sensory features may reveal distinct neurophysiological patterns and guide more personalized interventions.

## Conclusions

This study provides compelling evidence that migraine is characterized by altered brain signal complexity, with widespread reductions in entropy during resting state, especially in chronic migraine cases. These reductions reflect constrained neural processing and less adaptability in regions supporting visual processing, attentional control, self-referential processing, and cognitive and emotional regulation of pain. Importantly, we observed that migraine attacks are associated with transient increases in entropy that were characterized by elevated LLE, confirming their origin in weakly chaotic neural dynamics rather than purely random noise. This suggests a potential compensatory release from rigid neural states through a deterministic chaotic destabilization. Symptom-specific variations in entropy further emphasize the relevance of subjective experiences in shaping brain signal complexity in migraine. Together, our findings highlight the utility of complexity analysis for capturing dynamic brain alterations in migraine and point toward future directions for personalized interventions.

## Data Availability

The datasets generated and analyzed during the current study are not publicly available due to participant confidentiality and institutional regulations but are available from the corresponding author on reasonable request.

## Declarations

## Acknowledgment

We gratefully acknowledge Jacqueline Dobson and Ayesha Kousar for their invaluable contributions in project coordination, patient recruitment, and imaging support.

This work is dedicated to the memory of Safoura Jafari, a talented young mathematics student devoted to voluntary scientific work, with the aspiration of becoming a great scientist, whose passion for science and commitment to freedom continue to inspire.

## Ethics Approval and Consent to Participate

The study was approved by the Institutional Review Board of the University of Michigan Medical School. All participants provided written informed consent prior to participation.

## Authors’ Contributions

Majid Saberi contributed to the conceptualization, data curation, formal analysis, investigation, methodology, software development, validation, visualization, and writing of both the original draft and subsequent review and editing of the manuscript. Dajung J. Kim contributed to the investigation, validation, and to the review and editing of the manuscript. Xiao-Su Hu contributed to software development and to the review and editing of the manuscript. Alexandre F. DaSilva contributed to the conceptualization, methodology, investigation, supervision, project administration, funding acquisition, resources, validation, and to the review and editing of the manuscript.

## Declaration of generative AI and AI-assisted technologies in the writing process

During the preparation of this work, the authors used ChatGPT (OpenAI) in order to improve the readability and language of the manuscript. After using this tool, the authors reviewed and edited the content as needed and take full responsibility for the content of the published article.

