## Supplementary Material for "Reduced brain entropy in migraine with partial restoration during attacks: a resting-state fMRI study"

### Title

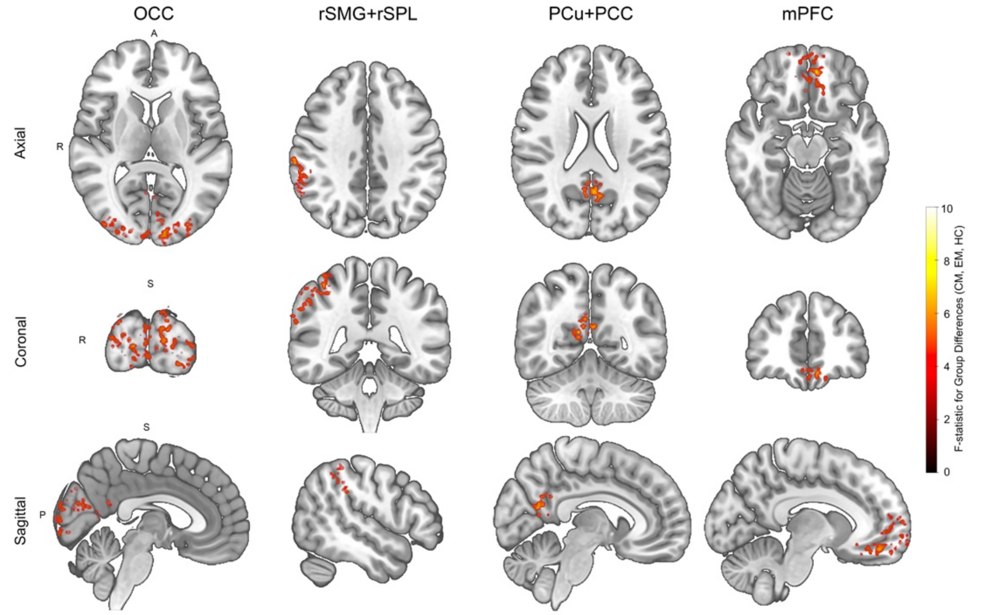


**Supplementary Figure S1. Nonparametric validation of group-level brain entropy differences using the Freedman-Lane permutation approach.** Brain maps display voxels exhibiting significant group-level differences in signal entropy among chronic migraine (CM), episodic migraine (EM), and healthy control (HC) groups based on a nonparametric permutation-based analysis of covariance (5,000 permutations), controlling for age and sex. The color scale represents permutation-derived F-statistics. Significant voxels surviving correction for multiple comparisons (exceeding 211 contiguous voxels) were clustered into four regions of interest: occipital cortex (OC), right supramarginal gyrus and right superior parietal lobule (rSMG+rSPL), precuneus and posterior cingulate cortex (PCu+PCC), and medial prefrontal cortex (mPFC), shown across axial, coronal, and sagittal views. The spatial distribution of significant clusters closely parallels the parametric results presented in Figure 1, supporting the robustness of the observed group differences.

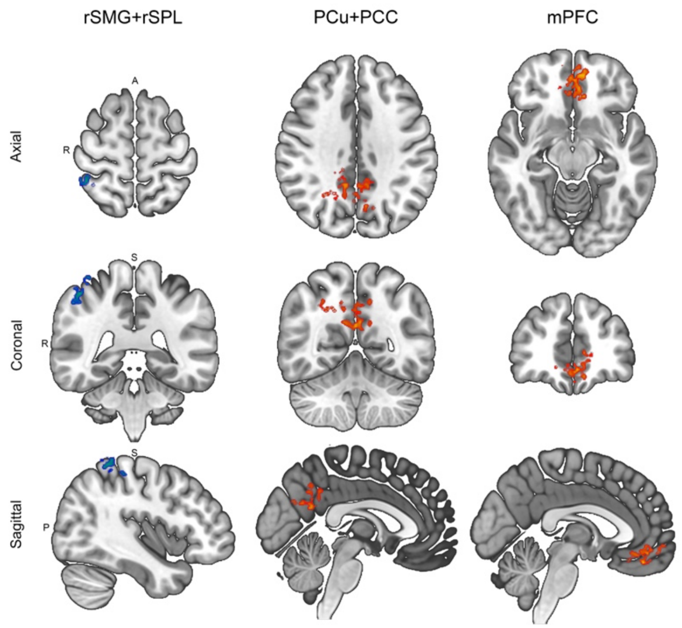


**Supplementary Figure S2. Sensitivity of group-level brain entropy results to alternative denoising strategy.** Brain maps display group-level differences in signal entropy among chronic migraine (CM), episodic migraine (EM), and healthy control (HC) groups following preprocessing with ICA-AROMA for denoising. Clusters surviving cluster-level correction for multiple comparisons are shown in warm colors and are localized primarily to the precuneus/posterior cingulate cortex (PCu+PCC) and medial prefrontal cortex (mPFC). Spatially consistent but subthreshold clusters (smaller than 209 voxels) within multisensory parietal regions encompassing the right supramarginal gyrus and superior parietal lobule (rSMG+rSPL) are shown in blue for illustrative comparison.

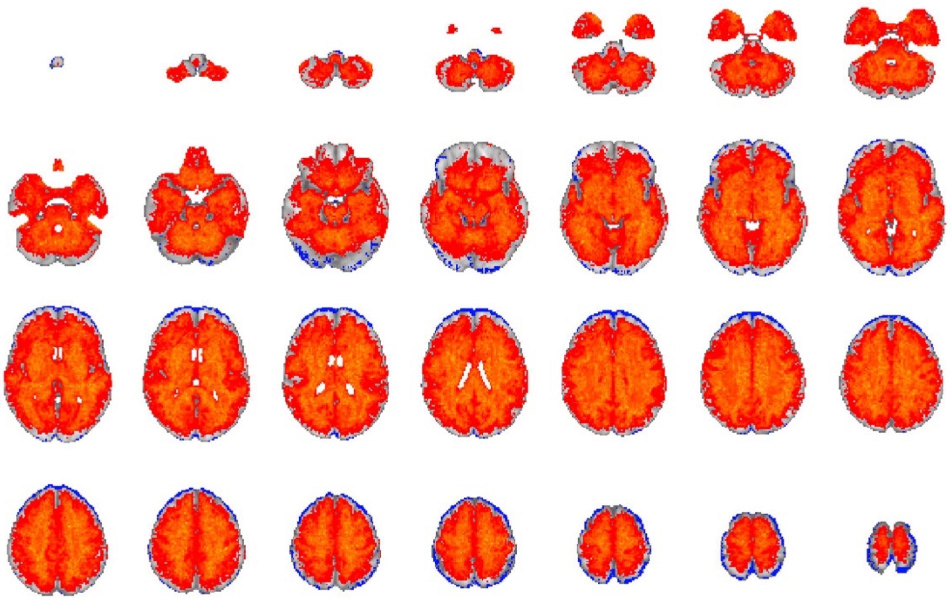

**Supplementary Figure S3. Voxelwise comparison of signal-to-noise ratio (SNR) between denoising pipelines.** Whole-brain maps depict voxelwise group-level differences in SNR following preprocessing with (i) aCompCor combined with regression of six motion parameters and (ii) ICA-AROMA. Red voxels indicate regions where the aCompCor + motion regression pipeline yielded significantly higher SNR, while blue voxels indicate regions where ICA-AROMA resulted in significantly higher SNR. Statistical comparisons were performed at the group level with correction for multiple comparisons. The spatial distribution demonstrates that the primary preprocessing pipeline (aCompCor + motion regression) produced higher SNR across the majority of brain regions, suggesting improved signal robustness relative to ICA-AROMA.

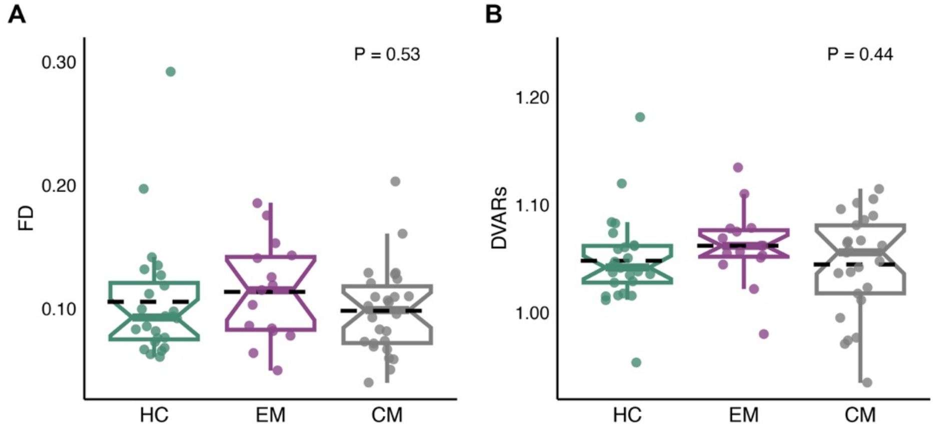


**Supplementary Figure S4. Group comparison of head motion metrics.** (A) Mean framewise displacement (FD) and (B) DVARS across healthy controls (HC), episodic migraine (EM), and chronic migraine (CM). No significant group differences were observed for either metric (FD: $p = 0.54$, $F = 0.816$; DVARS: $p = 0.45$, $F = 0.615$), indicating comparable head motion across groups.

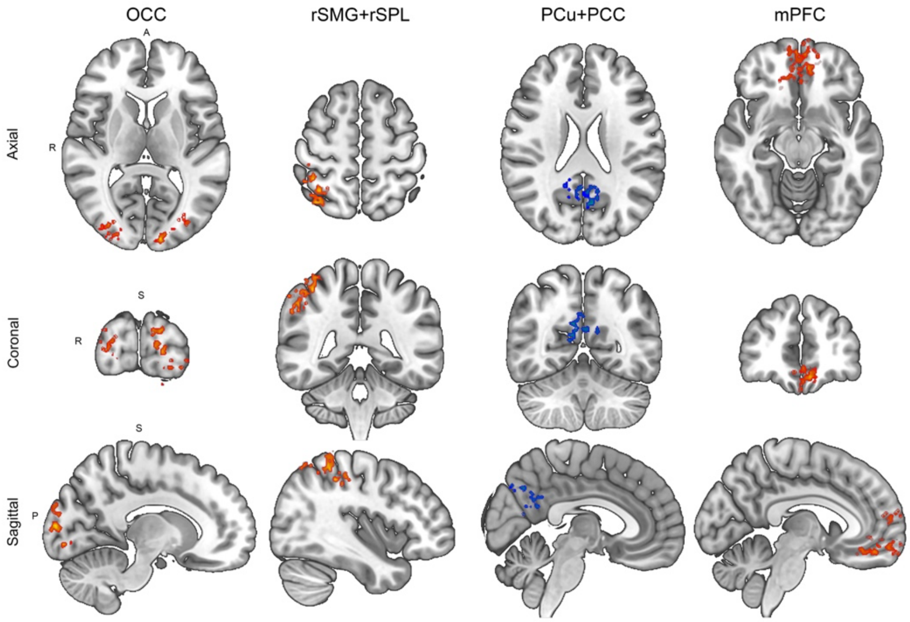


**Supplementary Figure S5. Sensitivity of group-level brain entropy results to embedding dimension (m = 3).** Brain maps display voxelwise group-level differences in signal entropy among chronic migraine (CM), episodic migraine (EM), and healthy control (HC) groups following recalculation of sample entropy using an embedding dimension of m = 3 while maintaining similarity tolerance at r = 0.2 × SD. Results are shown across axial, coronal, and sagittal views. Significant clusters surviving cluster-level family-wise error (FWE) correction are observed in the occipital cortex (OCC), right supramarginal gyrus and superior parietal lobule (rSMG+rSPL), and medial prefrontal cortex (mPFC). Although significant voxels were present within the precuneus/posterior cingulate cortex (PCu+PCC), this region did not reach the FWE cluster-size threshold (366 voxels) and is displayed in blue. Overall, the primary spatial pattern of entropy reduction across migraine groups is largely preserved, indicating robustness to changes in embedding dimension.

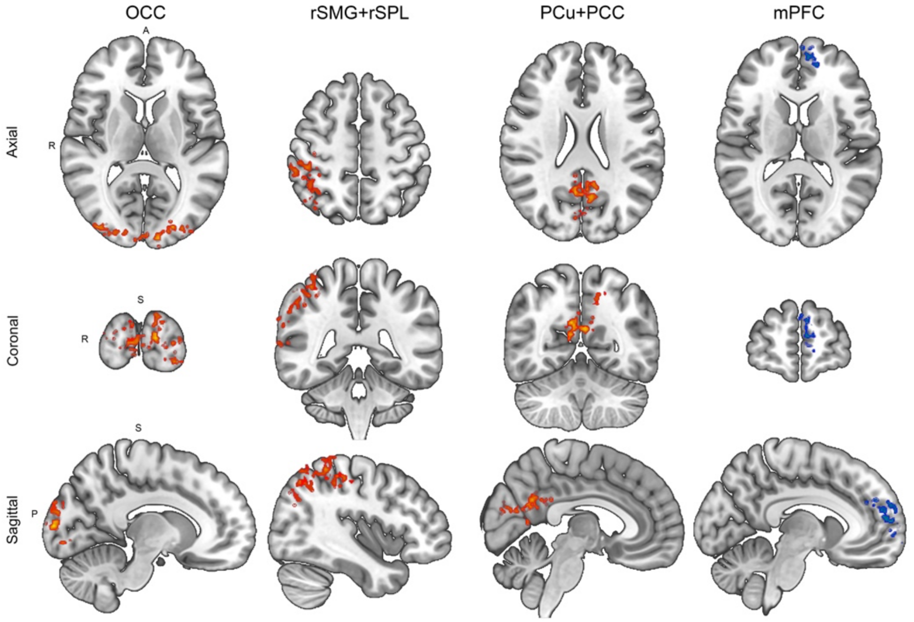


**Supplementary Figure S6. Sensitivity of group-level brain entropy results to similarity tolerance (r = 0.25 × SD).** Brain maps show voxelwise group-level entropy differences among chronic migraine (CM), episodic migraine (EM), and healthy control (HC) groups following recalculation of sample entropy using a similarity tolerance of r = 0.25 × SD while maintaining embedding dimension at m = 2. Significant clusters surviving cluster-level family-wise error (FWE) correction are observed in the occipital cortex (OCC), right supramarginal gyrus and superior parietal lobule (rSMG+rSPL), and precuneus/posterior cingulate cortex (PCu+PCC). Although significant voxels were present in the medial prefrontal cortex (mPFC), this region did not meet the FWE cluster-size threshold (337 voxels) and is displayed in blue. The replication of major clusters across parameter settings supports the stability of the main entropy findings.

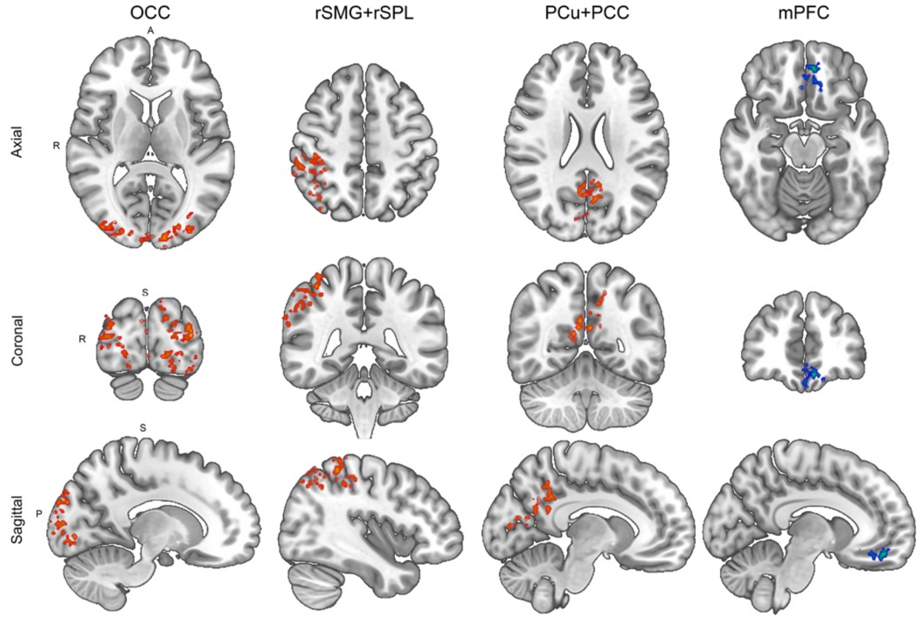


**Supplementary Figure S7. Sensitivity of group-level brain entropy results to inclusion of depressive symptom severity (BDI) as a covariate.** Brain maps display voxelwise group-level differences in signal entropy among chronic migraine (CM), episodic migraine (EM), and healthy control (HC) groups after including Beck Depression Inventory (BDI) score as an additional covariate alongside age and sex in the voxel-wise ANCOVA model. Significant clusters surviving cluster-level family-wise error (FWE) correction are observed in the occipital cortex (OC), right supramarginal gyrus and superior parietal lobule (rSMG+rSPL), and precuneus/posterior cingulate cortex (PCu+PCC), shown across axial, coronal, and sagittal views. Spatially consistent but subthreshold clusters within the medial prefrontal cortex (mPFC), which did not meet the corrected cluster-size threshold (207 contiguous voxels), are displayed in blue for illustrative comparison. The overall preservation of major clusters following inclusion of BDI as a covariate demonstrates that the principal entropy alterations are largely robust to adjustment for depressive symptom severity.
